# Participation after childhood stroke: Is there a relationship with lesion size, motor function and manual ability?

**DOI:** 10.1101/2021.03.22.21253256

**Authors:** Cristina Simon-Martinez, Sandeep Kamal, Fabienne Frickmann, Leonie Steiner, Nedelina Slavova, Regula Everts, Maja Steinlin, Sebastian Grunt

## Abstract

**Background:** Childhood arterial ischemic stroke (AIS) is associated with significant morbidity with up to 50% of affected children developing hemiparesis. Hemiparesis is assumed to influence participation within the peer group, but it is unclear to what extent its severity affects participation in different areas of social life.

**Methods:** Thirteen children (mean age 9y6m) with AIS (6 without hemiparesis, 7 with hemiparesis) and 21 controls (mean age 9y8m) participated. We scored hemiparesis severity with hand strength asymmetry (pinch and grip strength), measured with a dynamometer. We assessed manual ability (ABILHAND-Kids), socioeconomic status (Family Affluence Scale) and participation (Participation and Environment Measure – Children and Youth). From structural MRI, we measured lesion size. We investigated differences in participation and its relationship with hemiparesis severity using non-parametric partial correlations (controlling for lesion size, manual ability, and socioeconomic status), interpreted as absent (r<0.25), weak (r=0.25-0.50), moderate (r=0.50-0.75) or strong (r>0.75). Analyses were performed in jamovi 1.6.3.

**Results:** Children with AIS (with or without hemiparesis) showed reduced participation frequency at school (p<0.001), whilst participation at home and in the community resembled that of their peers. Severity of hemiparesis was moderately related to frequency and involvement at home and to involvement and desire for change in the community, although unrelated to school participation.

**Conclusion:** Reduced participation in school life requires close attention in the follow-up of children with AIS - regardless of the severity of hemiparesis. Participation at home and in the community is related to hemiparesis severity and may be improved with participation-focused motor intervention strategies.

**Highlights:**

- Children with stroke participate less frequently at school but are equally involved.
- Decreased participation at school is not related to hemiparesis severity.
- Participation at home and in the community are related to hemiparesis severity.

## 1. Introduction

Meeting friends, socializing, or doing sports are activities that we normally like doing and are part of our routine in different life situations (i.e., home, work, and community). The International Classification of Functioning, Disability and Health for Children and Youth (ICF-CY) defines participation as involvement in such life situations ^1^. Participation is, however, not defined as what a person can or wants to do. Children typically participate with high frequency and involvement in a variety of activities. Participation is an important activity in our lives, as it is crucial for the development of an identity, but also to become active, independent members of the society. The level of participation may be hampered by an event such as a stroke during childhood.

Arterial ischemic stroke (AIS) is a rare but devastating disease that may occur during childhood and may result in hemiparesis in up to 50% of children ^2–5^. Hemiparesis refers to the deficits (e.g., increased muscle weakness and spasticity, impaired range of motion, poor grasping and releasing skills) in one side of the body, contralateral to the hemisphere where the stroke occurred. These motor deficits may affect participation and quality of life ^6,7^ in children with AIS.

Participation-based studies in children with AIS are scarce but have emerged in recent years ^8^. Available studies identify reduced participation and social interaction with moderate difficulty, with some children having low or no such difficulties ^9,10^. Strikingly, poor participation in the community has been related to reduced quality of life in children with developmental disabilities ^11^. Despite a poorer participation in individual physical activities, team sports and cycling, research shows that children with motor impairment (resulting from AIS or other aetiologies) enjoy the activities as much as typically developing peers ^12^, when they participate in such activities. Studies have also reported that children with AIS have reduced social acceptance and social support from their peers ^13^ and they often change friendships ^14^. As the clinical presentation of AIS is very heterogeneous, some studies have focused on clinical subgroups. Greenham et al (2018) recently reported no differences in participation between subgroups of children with AIS (neonatal (<29 days) or childhood-onset (>29 days)) using the Participation and Environment Measure - Children and Youth (PEM-CY) questionnaire ^15^. Given this evidence and that not all the children with AIS will have hemiparesis, it is important to further investigate restrictions in participation between children with AIS, and to what extent such restrictions are related to hemiparesis.

In the ICF-CY, in addition to the domain of participation, a distinction is made between body structures and body functions, activity, person-related factors and environmental factors. These domains interact with each other and influence the individual’s health condition. Whilst the relationship between these components and participation has been previously studied in children with cerebral palsy (irrespective of the type of brain lesion) ^16–20^, there is a limited number of studies in children with AIS. Participation restriction was shown to be linked to subcortical AIS (*body structures*) ^10^, impairments in gross motor function (*body functions*) ^15^, and limitations in manual ability (*activity*) ^21^. Additionally, a key predictor of participation at home and in the community in children with AIS is fatigue (*body functions*) ^15^. Impaired cognition and more behavioural problems (cognition and behaviour, *body functions*) also positively correlate with restrictions in participation ^21^.

Environmental factors are defined as “the physical, social and attitudinal environment in which people live and conduct their lives” ^1^. They have a crucial role in how a child participates. The involvement of the child in different life situations is very much linked to the family environment, which is integral to understanding participation in early childhood ^1^. The changes in these environmental factors may have an impact on the person’s competencies and independence. Whilst there is no specific evidence for the interaction between environmental factors and participation in children with AIS, there is evidence showing that higher levels of family cohesion, higher incomes, better family coping strategies, and lower levels of stress may facilitate participation in children and adolescents with cerebral palsy ^16,17^.

Overall, there are some hints on what participation in children with AIS looks like, although much remains unknown. It remains unclear how a hemiparesis after AIS influences the participation pattern and to what extent participation in children with AIS is related to different components of the ICF-CY. To develop new treatment strategies to improve participation in children diagnosed with AIS, their participation restrictions and the factors that influence them need to be better understood. Therefore, the aim of this study was threefold. Our **first aim** was to identify differences in participation patterns in various life situations (home, school, and community) between children diagnosed with AIS with and without hemiparesis and typically developing peers. The **second aim** was to explore the relationship between participation restriction and the different ICF-CY components in an AIS cohort. Lastly, the **third aim** was to examine the unique relation between participation restriction and severity of hemiparesis controlling for the other influencing ICF-CY components (lesion size, manual ability and socioeconomic status).

## 2. Materials and Methods

### 2.1. Participants

Children and adolescents with and without AIS were recruited as part of the ‘Hemispheric Reorganization (HERO)’ study ^22^ that investigated cortical reorganization after paediatric AIS. The participants with AIS were recruited from the population-based Swiss Neuropediatric Stroke Registry ^2^. Children and adolescents of the AIS group were *included* if: (i) they had an AIS before the age of 16 years (confirmed by MRI or computed tomography) at a chronic stage (i.e., at least two years prior to the study), (ii) an age of at least 5 years old at the time of assessment. *Exclusion* criteria were (i) those related to MR compatibility, (ii) concomitant neurological disorders and (iii) behavioural problems that would interfere in the assessment. Of the 283 patients from the Swiss Neuropediatric Stroke Registry who met the eligibility criteria and were contacted, a final sample of 28 patients agreed to participate in the HERO study (more details about the reasons for not participating can be found in the original publication of the HERO study) ^23^. A control group of typically developing peers was recruited through advertisement in the university hospital intranet and flyers. The inclusion criterion was the absence of any developmental disorder, the exclusion criteria were the same as for the AIS group. Two years after the initial assessment, participants of the HERO study were contacted and asked to complete an additional questionnaire regarding participation and socioeconomic status. For the questionnaire, only native German speaking families whose children were younger than 17 years old at the time of completing the questionnaire were contacted. The final study population of the present sub-study consisted of 13 children diagnosed with AIS and 21 typically developing peers. The Research Ethics Committee of Berne, Switzerland approved the SNPSR, the HERO study, and the present study. All parents and legal guardians provided written consent and the participants assented to partake in the study, in accordance with the Declaration of Helsinki.

### Evaluation

We gathered demographic and other basic data (sex, age at assessment, age at stroke and time since stroke) from the SNPSR and at the time of the first assessment. Handedness was determined at the first assessment (thus in the chronic stage after AIS) with the Edinburgh Handedness Inventory^24^.

For the assessment of childhood AIS at all the ICF-CY levels we included brain lesion size (body structure), hand strength (body function, motor), manual ability (activity), participation (participation patterns of children at home, school and in the community) and socioeconomic status and parents’ educational level (environment) (Figure 1).

**Figure 1.**
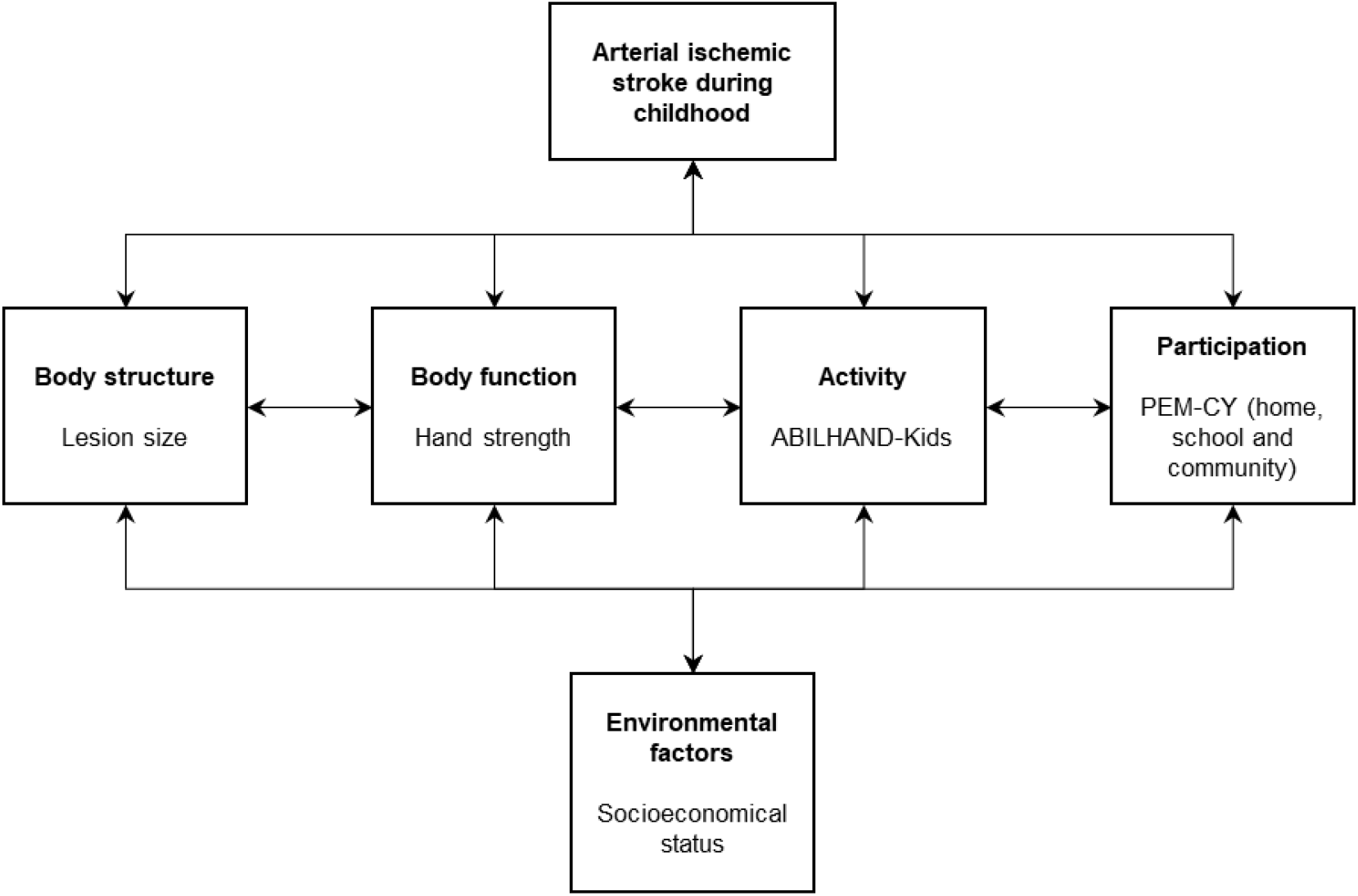
Scheme of the International Classification of Functioning (Children and Youth version) and the outcome measures used in this study.

At the **body structure level**, we included *lesion size*. Lesion size was measured with the ABC/2 method. The exact procedure for calculating the lesion size was described previously ^25,26^. Briefly, lesions were marked in the structural T1 images and their volume was calculated based on these markings. The volume of the lesion was divided by the total intracranial volume and expressed in mm^3^. Additionally, we extracted the *location and side of the lesion* for completeness of the database, although these data were not used for statistical analyses. Lesions were classified according to their location: cortical, subcortical, and combined cortical and subcortical. Lesion laterality was categorized into left, right, and bilateral.

At the **body function level**, we included *hand strength*. Hand pinch and grip strength was used as an index for the body function impairments after AIS that would quantify the severity of hemiparesis motor impairment. Hand strength was evaluated with a dynamometer (30 psi pneumatic dynamometer Baseline®, USA and a 30 lb mechanical pinch gauge, Baseline®, USA). First, the maximum voluntary contraction of each hand was recorded three times and the average value was retained. Next, we calculated the mean of the pinch and grip average values. The asymmetry index to quantify the severity of hemiparesis was computed as the percent of asymmetry of hand strength between the dominant and the non-dominant side, as follows:

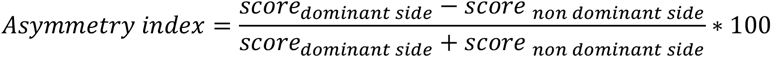

A higher score indicates larger asymmetry toward the dominant side, whilst a score of 0 indicates symmetric strength between both hands.

At the **activity level**, we included *manual ability*, evaluated with the ABILHAND-Kids, which is a valid and reliable measure to assess manual ability through the scoring of 21 daily activities. The difficulty experienced by the child to perform the required tasks is rated on a 3-point ordinal scale by the parents 27.

We measured **participation** with the Participation and Environment Measure for Children and Youth (PEM-CY). This questionnaire examines participation frequency, the extent of involvement, and desire for change in sets of activities typical for the home, school, or community ^28,29^. Participation *frequency* in percentage provides an indicator of the frequency of engagement in activities (higher scores indicate greater frequency). Participation *involvement* indicates the extent to which a child is engaged in the activities, including social engagement (higher scores indicate greater involvement). The *percent of activities* indicates the range of activities the child participates in (higher scores indicate greater number of activities). The *desire for change* indicates the satisfaction with the current participation (higher scores indicate greater desire for change).

To evaluate the impact of **environmental** factors influencing participation, we included measures of *socioeconomic status* (SES) by using the Family Affluence Scale. This scale is commonly used to identify the SES of children and adolescents in studies on child health and well-being ^30^. It includes four questions that reflect the privileges that the family may have. Additionally, we included questions about the *parents’ educational level* (one for each parent). The used questions can be found in Appendices (Document A.1). The final scale score ranged from 0 to 15, with higher values indicating more privileged families with higher parental educational level.

### 2.2. Statistical analyses

Due to the low number of participants, we opted to perform non-parametric statistics. For the **first aim** (*what are the differences in participation between groups?*), we first divided the AIS cohort into those who developed a hemiparesis and those who did not. For that, we used the motor sub-score of the Paediatric Stroke Outcome Measure and those who scored 0 were labelled as AIS without hemiparesis and those who scored more than 0 were labelled as AIS with hemiparesis. To investigate differences in participation characteristics between the three groups, we conducted the Kruskal-Wallis test and the respective pairwise comparisons with the Dwass-Steel-Critchlow-Fligner test ^31^. The level of significance was set at α=0.05. To report effect sizes from the Kruskal-Wallis tests, we used epsilon square (ε^2^) ^32^. An ε^2^ of 0 would mean no differences and no influence, while 1 would indicate a full dependency. We interpreted effect sizes as none (<0.25), weak (0.25-0.50), moderate (0.50-0.75) or strong (>0.75) ^32^.

The second and third aims focused on the AIS cohorts. For the **second aim** (*what is the relation between severity of hemiparesis (hand strength asymmetry) and participation?*), we used non-parametric correlations to document the correlation between each pair of variables included in the study. To answer our **third aim** (*what is the unique relation between hemiparesis severity and participation?*), we ran non-parametric partial correlations, as they allow for the elimination or control of possible influencing variables, to investigate the unique relation between the severity of hemiparesis and participation, controlling for lesion size, manual ability, and SES. All correlations were reported with Spearman’s rho and interpreted as absent (<0.25), weak (0.25-0.50), moderate (0.50-0.75) or strong (>0.75). The level of significance was set at α=0.05.

All analyses were conducted with jamovi for Windows v1.6.3.

## 3. Results

### 3.1. Participants

Thirty-four children participated in the study: 13 with AIS and 21 controls. In the AIS cohort, there were 6 children without hemiparesis and 7 with hemiparesis. The three cohorts were similar in age and sex, and there were more right-handed participants in the control cohort (Table 1). Between the AIS cohorts (with and without hemiparesis), there was no difference in lesion side or location, but lesion size was larger in the cohort with hemiparesis (p=0.03, Table 1).

**Table 1.** Demographic characteristics of each cohort and group comparison.

|  |  | Controls<br>(n=21) | AIS without<br>hemiparesis<br>(n=6) | AIS with<br>hemiparesis<br>(n=7) | p |
| --- | --- | --- | --- | --- | --- |
| Age at motor eval. | Mdn (IQR) | 9.8 (7.2-10.8) | 10.1 (8.6-11.3) | 9.5 (9.1-12.3) | 0.90 |
| Age at PEM-CY eval. | Mdn (IQR) | 12.0 (9.0-13.0) | 13.0 (12.0-14.0) | 13.0 (12.0-15.5) | 0.39 |
| Age at stroke | Mdn (IQR) |  | 2.4 (0.8-4.6) | 1.5 (0.8-4.4) | 0.77 |
| Sex | Female (n (%)) | 8 (38) | 2 (33.3) | 2 (28.6) | 0.90 |
|  | Male (n (%)) | 13 (62) | 4 (66.7) | 5 (71.4) |  |
| Handedness | Left-handed (n (%)) | 2 (10) | 0 (0.0) | 4 (57) | 0.01* |
|  | Right-handed (n (%)) | 19 (90) | 6 (100.0) | 3 (43) |  |
| Lesion Side | Left (n (%)) |  | 5 (83) | 6 (86) | 1.00 |
|  | Right (n (%)) |  | 1 (17) | 1 (14) |  |
| Lesion Size | Mdn (IQR) |  | 0.0 (0.0-0.6) | 2.6 (1.8-3.2) | 0.03* |
| Lesion Location | Cortical (n (%)) |  | 3 (50.0) | 1 (14.3) | 0.08 |
|  | Subcortical (n (%)) |  | 3 (50.0) | 2 (28.6) |  |
|  | Combined (n (%)) |  | 0 (0.0) | 4 (57.1) |  |

Eval., evaluation; Mdn, Median; IQR, interquartile range; n, number of participants; PEM-CY, Participation and Environment Measure - Children and Youth.

### 3.2. Aim 1 - Differences in participation and other components of the ICF-CY between AIS and control cohorts

We compared the outcomes at all levels of the ICF-CY between the control cohort and each of the AIS cohorts with and without hemiparesis and a summary of this comparison can be found in Table 2. We found differences in **lesion size** (body structure, between AIS cohorts) whereby those with hemiparesis had a larger lesion (p=0.03). The **hand strength asymmetry index** (body motor function) was more imbalanced in the AIS with hemiparesis cohort (p=0.02). Post-hoc analyses depicted a trend of more asymmetry toward the dominant hand in the AIS with hemiparesis cohort, compared to controls (p=0.08). At the activity level, we found group differences in the ABILHAND-Kids (p<0.001) that reflected poor **manual ability** in the AIS cohorts compared to the control cohort (post-hoc analyses control vs. AIS (with or without hemiparesis), p<0.01), whereas manual ability did not relate to hemiparesis or not.

**Table 2.** Descriptive statistics (median and interquartile range) of the outcome measures at each level of the ICF-CY in each cohort and group comparison.

| | N | Controls (n=21) | AIS without hemiparesis (n=6) | AIS with hemiparesis (n=7) | p ($\epsilon^2$) |
| --- | --- | --- | --- | --- | --- |
| **Body structure** |  |  |  |  |  |
| Lesion size | 13 | - | 0.0 (0.0-0.6) | 2.6 (1.8-3.2) | 0.03* (0.38) |
| **Body function** |  |  |  |  |  |
| Hand strength asymm. | 34 | 1.5 (-1.6-3.7) | -3.2 (-4.1--2.5) | 18.9 (6.1-38.9) | 0.02*<sup>¥</sup> (0.24) |
| **Activity** |  |  |  |  |  |
| ABILHAND-Kids (logits) | 34 | 6.7 (6.7-6.7) | 4.4 (3.9-5.9) | 3.2 (2.2-5.0) | <0.001*<sup>§</sup><sup>¥</sup> (0.54) |
| **PEM-CY Home** |  |  |  |  |  |
| Frequency (%) | 34 | 87.1 (78.6-90) | 87.1 (82.5-89.6) | 81.4 (77.1-83.6) | 0.58 (0.01) |
| Involvement | 33 | 4.3 (4.2-4.5) | 4.2 (4.0-4.4) | 3.9 (3.8-4.4) | 0.17 (0.06) |
| Activities (%) | 34 | 100 (100-100) | 100 (100-100) | 100 (90-100) | 0.65 (0.01) |
| Desire for change (%) | 33 | 40 (30-50) | 60 (30-60) | 50 (30-55) | 0.40 (0.02) |
| **PEM-CY School** |  |  |  |  |  |
| Frequency (%) | 34 | 58.6 (50.7-67.1) | 25 (21.8-27.1) | 22.9 (19.3-24.3) | <0.001*<sup>§</sup><sup>¥</sup> (0.72) |
| Involvement | 33 | 4.7 (4.2-5.0) | 4.2 (3.6-4.7) | 4.5 (4.1-5.0) | 0.35 (0.03) |
| Activities (%) | 34 | 80 (80-100) | 70 (60-80) | 60 (60-70) | 0.04*<sup>¥</sup> (0.13) |
| Desire for change (%) | 33 | 0.0 (0.0-20.0) | 20.0 (0.0-20.0) | 20.0 (20.0-87.5) | 0.05* (0.13) |
| **PEM-CY Community** |  |  |  |  |  |
| Frequency (%) | 34 | 42.9 (32.9-60.4) | 36.4 (23.6-45) | 41.4 (32.1-44.3) | 0.11 (0.08) |
| Involvement | 33 | 4.7 (4.1-5.0) | 4.2 (3.9-4.4) | 4.5 (4.2-4.9) | 0.62 (0.01) |
| Activities (%) | 34 | 80.0 (70.0-90.0) | 80.0 (50.0-80.0) | 70.0 (60.0-75.0) | 0.20 (0.05) |
| Desire for change (%) | 33 | 10.0 (0.0-20.0) | 20.0 (0.0-30.0) | 30.0 (5.6-40.0) | 0.12 (0.07) |
| SES | 34 | 13.0 (12.0-15.0) | 12.0 (9.2-14.8) | 14.0 (11.5-16.5) | 0.73 (0.02) |

AIS, Arterial Ischemic Stroke; n, number of participants; PEM-CY, Participation and Environment Measure - Children and Youth; $\epsilon^2$ , epsilon square (measure of effect size). Post-hoc analyses: <sup>¥</sup>control vs. AIS with hemiparesis, <sup>§</sup>control vs. AIS without hemiparesis.

At the level of **participation at home**, we found no differences between cohorts (p>0.05) and the effect sizes were small (ε^2^<0.10). **Participation at school** was different between cohorts (p<0.001) with a moderate-large effect size (Table 2). Post-hoc analyses indicated that children with AIS (with or without hemiparesis) participated less frequently at school (p<0.001) compared to controls. Also, the percentage of activities in which they were involved at school was different (p=0.04) and post-hoc analyses showed a trend toward significantly different scores between control and AIS with hemiparesis (p=0.06). Similarly, the desire for changing their participation at school was rated higher for the AIS cohorts (p=0.05), although it was not significant in post-hoc analyses. **Participation in the community** did not differ between cohorts (p>0.05) and the effect sizes were small. Figure 2 depicts the responses of the PEM-CY questionnaire per group and per domain and individual data of the used variables can be found in Appendices (Figure A.1).

**Figure 2.**
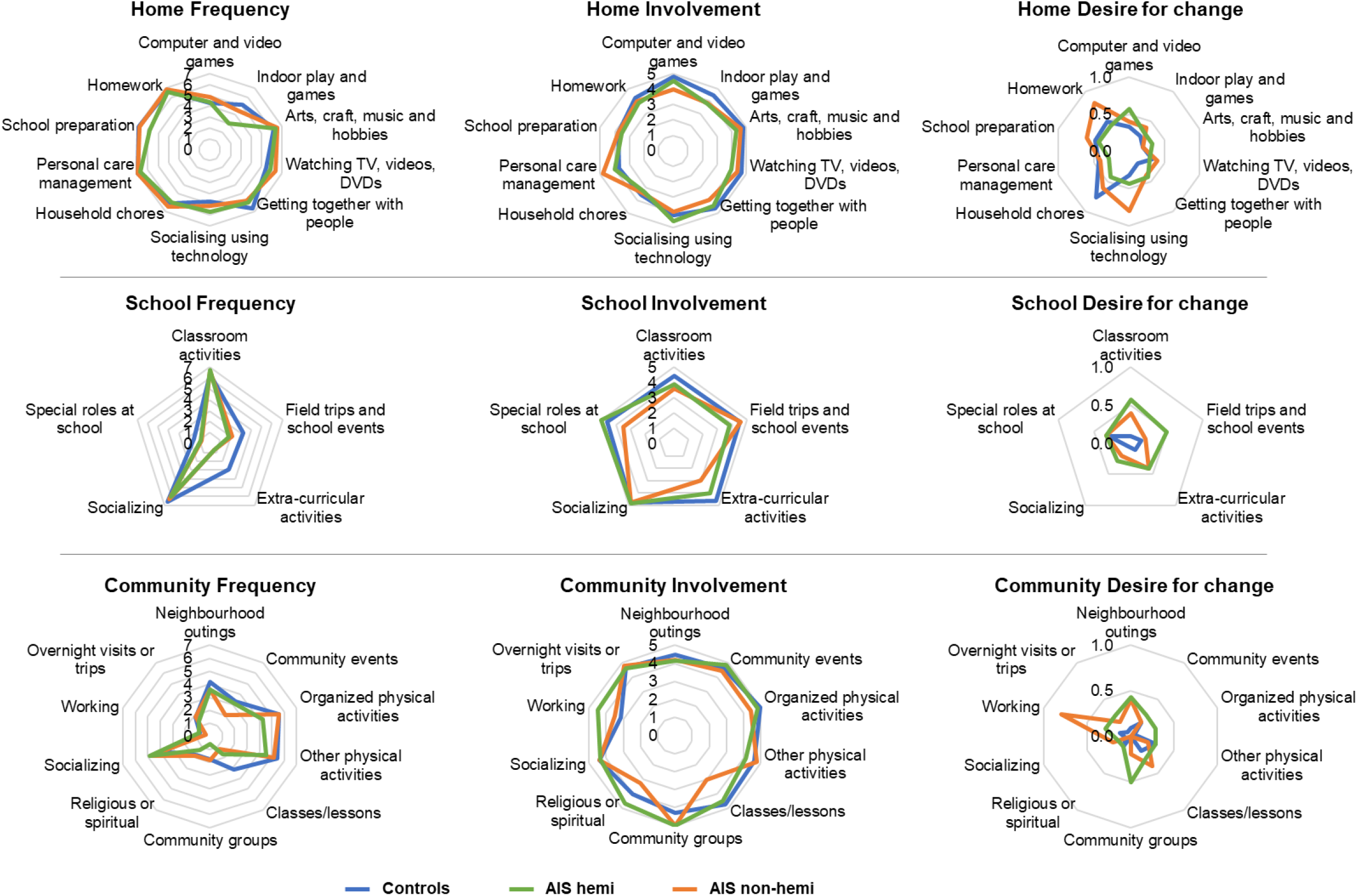
Differences in participation frequency, involvement, and desire to change (at home, school, and in the community) between the control and AIS with or without hemiparesis cohorts for every item of the PEM-CY questionnaire.

At the environment level, we found that the **SES** was not different between cohorts (p>0.05).

### 3.3. Aim 2 – Bivariate relation between participation and other measures at the ICF-CY levels

We described an overview of the relation (bivariate correlations) between the different levels of the ICF-CY and the measures of participation in Figure 3. We found weak negative relations between **lesion size** and frequency of **participation** at home and school, whereby the larger the lesion the lower the participation frequency. Correlation analyses showed weak to moderate positive relations between lesion size and the involvement at home, school and in the community, suggesting that the larger the lesion size, the higher the involvement. The percent of activities in which the children participated also correlated negatively (weak to moderate) with lesion size, whereby the larger the lesion, the lower the percent of activities. Similarly, the desire for change the participation level at home and in the community negatively and moderately correlated with lesion size, suggesting that the parents would like a larger change in the participation at home and in the community with smaller lesion sizes.

**Figure 3.**
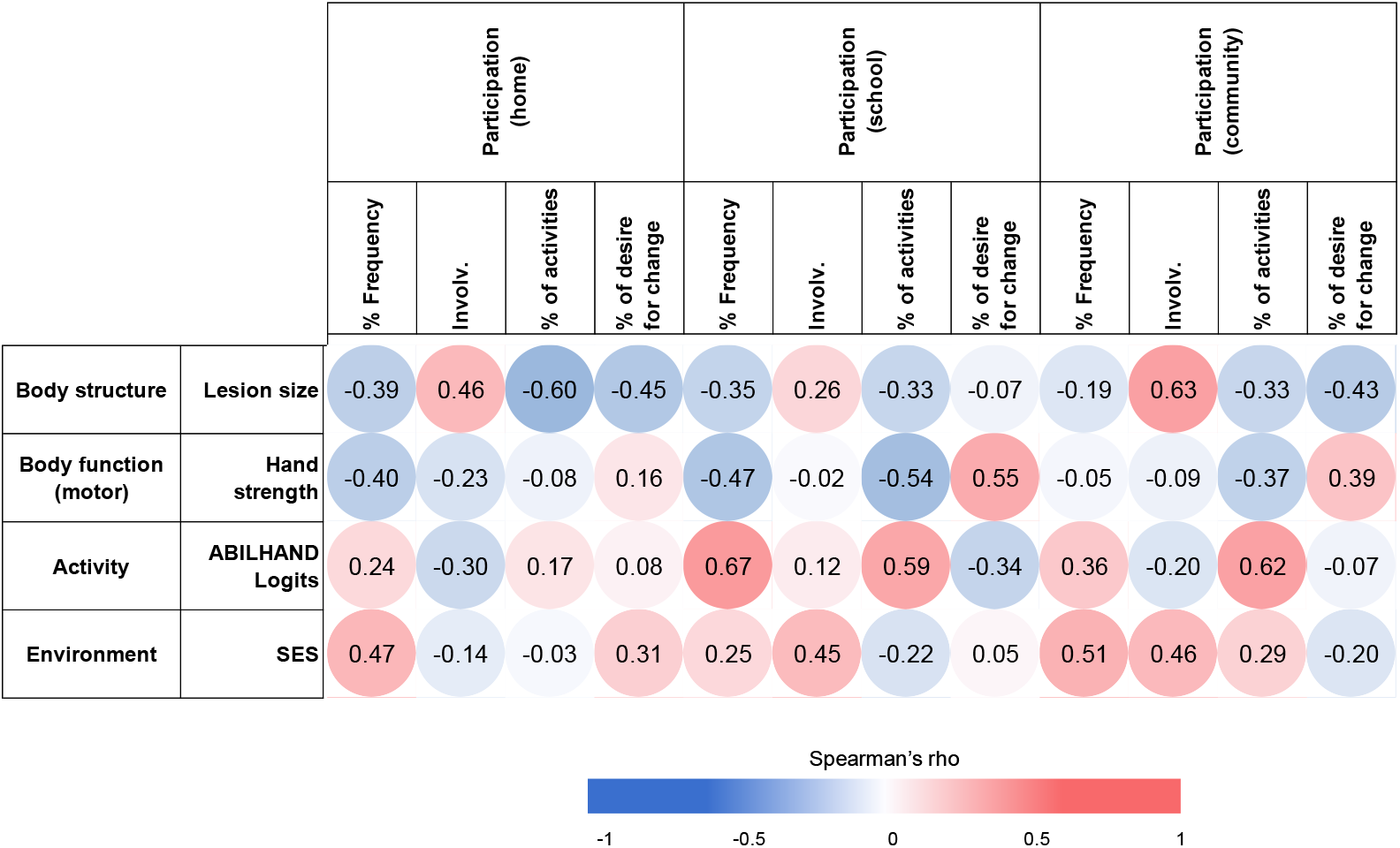
Correlation matrix between the different levels of the ICF model (body structure, body function, activity, participation, and environment) in the AIS (with or without hemiparesis) cohort.

**Hand strength** at the body function level was mostly correlated with **participation** at school. We found moderate negative correlations between hand strength asymmetry index and frequency of participation and percent of activities (Figure 3). These correlations indicated that the more affected hand strength was (higher values) the lower the participation at school. At school and in the community, we found a moderate positive correlation between hand strength and percent of desire for change, suggesting that with more impaired hand strength, parents increased their desire for the participation to change.

At the **activity** level, the ABILHAND-Kids positively correlated with **participation** measures at school and in the community. More specifically, we found positive moderate correlations between manual ability and frequency of participation and percent of activities, indicating that participation increased with better manual ability.

Lastly, the **environmental** measures positively correlated with **participation** measures at home, school and in the community, suggesting that the better the socioeconomic status and education level of the family, the higher the participation was, in terms of frequency of participation and involvement.

### 3.4. Aim 3 – Unique relation between hand strength asymmetry and participation measures

Overall, our study shows that even when controlling for other factors, the severity of hemiparesis is related to participation. More specifically, the partial correlation between hand strength asymmetry and participation frequency and involvement at *home* increased to moderate correlations (rho=-0.51- −0.64). Similarly, the correlations between hand strength asymmetry and participation involvement and desire to change in the *community* increased to moderate correlations (rho=-0.54-0.53), whilst the relation with percent of activities decreased to no correlation. Lastly, the partial correlation between hand strength asymmetry and participation frequency, percent of activities and desire to change at *school* decreased in strength to weak (rho=0.43) and no correlation (rho=-0.13- −0.20), putting forward the influence of the covariates in the relationship. More details about the results of the partial correlations can be found in Table A.1.

## 4. Discussion

This study is the first one investigating restrictions in participation in children with AIS (with or without hemiparesis) and its relationship with other components of the ICF-CY. The advantage of having a more homogeneous sample (diagnosis of AIS) allows us to draw more specific conclusions whilst it also helps clinicians in their decision making.

We first aimed to investigate differences in participation and other components of the ICF-CY between AIS and typically developing controls. We found that children with hemiparesis due to AIS had a larger lesion than those without hemiparesis and AIS. This result is in agreement with previous studies showing the relationship between outcome severity and lesion size ^33,34^. We also found that the hand strength asymmetry index was most impaired in the cohort of children with hemiparesis, which seems logical, whilst children with AIS without hemiparesis did not show such motor impairments ^35^. However, the manual ability was decreased in both groups of children with AIS (with and without hemiparesis). We measured manual ability with the ABILHAND-Kids, that evaluates the child’s ability to manage daily activities that require the use of the upper limbs, whatever the strategies involved ^27^. The latter is important, as it seems that children with AIS who do not develop a hemiparesis may also have difficulties in finding strategies to use their upper limbs efficiently to manage daily activities, which may also affect their quality of life ^7^. Children with AIS who do not develop hemiparesis may also suffer cerebral blood flow imbalance ^36^, which may affect the way they use their upper limbs for uni- and bimanual daily life activities. This finding is clinically relevant, as children with AIS without hemiparesis may benefit from physical or occupational therapy programs to increase the use of their upper limbs in daily life activities and, consequently, increase their quality of life.

Within the first aim, we also investigated deficits in participation between the three groups. Participation at home and in the community did not differ between groups and the effect sizes were small. Previous studies reported that children with AIS may participate less and have less social interactions than their peers ^9,10^. However, the use of various tests to evaluate participation may be important for the sensitivity of the questionnaire to detect differences. A recent study used the PEM-CY questionnaire and found no differences between children with different AIS onsets (neonatal vs. childhood) ^15^. A study with a larger sample size would be needed to confirm our findings.

Interestingly, we found that participation at school differed between groups. First, children with AIS (with or without hemiparesis) participated less frequently than their peers and those with hemiparesis participated also in fewer activities. Remarkably, children with AIS seemed as involved as their peers whenever they participated in the activities, which is in line with previous research ^12^. In general, parents of children with AIS and hemiparesis seemed to desire that the participation pattern changed, although this was not statistically significant in post-hoc analyses.

The fact that participation differed only at school and not at home or in the community calls for further reflection. At home and in the community, the child may participate more because the peers may be more understanding of the child’s condition and needs, which may facilitate participation in these settings. In contrast, bullying and similar experiences at school, which seem to happen often in children with cerebral palsy (due to AIS or other type of brain injury) ^37^, may hamper participation in these settings. Nevertheless, our findings in AIS disagree with studies including a large, although the heterogeneous, population of children with disabilities, which showed reduced participation in all settings for children with disabilities ^38^. Further research will warrant more conclusive answers to this regard.

Our second and third aim are interrelated, and as the second one is not completely novel, we will focus on discussing the results of the unique relation between the severity of hemiparesis and participation measures. Our main finding was that the severity of hemiparesis (measured with hand strength asymmetry) was moderately correlated with participation frequency and involvement at home, as well as participation involvement and desire for change in the community. Our results suggested that with more severe hemiparesis, the participation occurred with lower frequency and involvement, and the parents had higher desire for change that participation pattern. As we used partial correlations, these results show the unique relationship between the severity of hemiparesis and participation measures, controlled by other ICF-CY components (lesion size, manual ability, and socioeconomic status). Previous studies have shown that participation in children with disabilities could be related to lesion characteristics ^10^, manual ability and activity limitations ^16–21^, and socioeconomic status ^16,17^. Our study shows that even when controlling for these factors, the severity of hemiparesis is related to participation. This fact should be taken onto the rehabilitation process.

At school, we found a weak relation between the severity of hemiparesis and the desire for change, although it seemed to be influenced by other factors (lesion size, manual ability, and socioeconomic status). When looking at the bivariate correlations (Figure 3), we can see that participation at school (and in the community) is strongly related to manual ability. Manual ability was measured with the ABILHAND-Kids, which evaluates the capacity to perform daily activities with the upper limbs, whatever the strategies involved ^27^. It is plausible that the strategies used at school and in the community enhance participation in these settings, whilst participation at home may be more demanding due to potential parental suggestions to use more the impaired hand in different tasks. Lastly, frequency of participation in the community was not related to severity of hemiparesis in the partial correlations, whilst it was moderately correlated to socioeconomic status. Socioeconomic status may facilitate engagement in physical activities in the community, where they need to be more independent ^17^, likely due to costs of adapted sports programs and equipment. The socioeconomic status of the families should be considered when intervening on participation with the support of policymakers.

Our study may have some clinical implications. The ultimate goal of any rehabilitation program should be targeting changes in participation. Understanding what the determinants of participation are in children with AIS is crucial to coordinate a multidisciplinary and individualized treatment strategy. Our study shows that the severity of hemiparesis may be one of the main determinants of participation at home and in the community. We know that family-centered services promote the psychosocial well-being of children and their parents ^39^, which has an immediate impact on participation ^15^. Participation-focused rehabilitation programs, embedded in the family, that also influence physical activity and body functions may be an optimal way to improve participation while acting on the body functions in the long run. Nevertheless, we need to better understand what resources parents need to support these types of interventions ^40^ so that they are provided as part of their child’s rehabilitation process.

We can address some limitations while interpreting current study results. Whilst we aimed to have a homogenous sample (AIS diagnosis with and without hemiparesis), our sample size was limited and the results on the participation questionnaire very variable, which may call for replication in future studies. For the third aim, we chose for non-parametric partial correlations due to the small sample size, although a regression model would be more appropriate and would allow us to estimate the individual influence of each factor on the model. Lastly, whilst we attempted to cover the most common deficits in AIS with the ICF-CY, there are other aspects, such as independence, coping and body image or cognitive and social difficulties (as children with cognitive impairments may be at higher risk of developing problems in the relation with their peers ^41^). These aspects should be explored in the future when investigating participation.

## 5. Conclusion

Children with AIS show more difficulties in participating at school, whilst they participate similarly to their peers in other settings (home and community), regardless of the severity of their hemiparesis. Whilst several components of the ICF-CY (lesion size, manual ability, and socioeconomic status) are related to participation in children with AIS, the severity of hemiparesis seems to play a more important role in participation frequency and involvement at home and in the community, but not at school. These findings highlight the importance of developing participation-focused training strategies for children with AIS (with and without hemiparesis) that have an additional impact on body functions.

## Data Availability

Data is available upon reasonable request to the corresponding author.

## Acknowledgements

The authors thank all the children who participated in the study as well as their parents. Furthermore, a big thank you goes to all co-workers at the Swiss Neuropediatric Stroke Registry: Andrea Capone Mori (Aarau), Alexandre Datta (Basel), Joël Fluss (Geneva), Annette Hackenberg (Zürich), Elmar Keller (Chur), Oliver Maier (St. Gallen), Jean-Pierre Marcoz (Sion), Claudia Poloni (Lausanne), Gian Paolo Ramelli (Bellinzona), Maria Regenyi (Berne), Regula Schmid (Winterthur), and Thomas Schmitt-Mechelke (Lucerne). We would also like to thank Alain Kaelin-Lang for his scientific assistance and Salome Kornfeld and Juan Delgado Rodriguez for their numerous hours of help with clinical data acquisition. In addition, we thank Susan Kaplan for the English language polishing. This study was supported by the Swiss National Foundation (32003B_146894/1), the Anna Müller Grocholski Foundation (Switzerland), the Swiss Foundation for Children with Cerebral Palsy and the Foundation Vinetum (Switzerland). The sponsors had no role in data collection, analysis, or interpretation, nor in the writing of the report or in the decision to submit the article for publication.

## Data availability

Data is available upon reasonable request to the corresponding author.

## Appendices

**Document A.1**. Questions used in the questionnaire of Family Affluence Scale (questions 1-4) and parents’ educational level (questions 5-6). The numbers in brackets indicate the score given for each response.

1. Does your child have a room all to themselves?
  a. No (0)
  b. Yes (1)
2. Do you own a vehicle as a family (car, van or camper)?
  a. No (0)
  b. Yes, one (1)
  c. Yes, two (2)
  d. Yes, more than two (3)
3. How often did you go on holiday with your child in the last 12 months?
  a. Not at all (0)
  b. Once (1)
  c. Twice (2)
  d. More than twice (3)
4. How many computers do you own as a family?
  a. None (0)
  b. One (1)
  c. Two (2)
  d. More than two (3)
5. What degree does the mother have?
  a. No degree (0)
  b. Junior high school (intermediate school certificate) (1)
  a. Secondary school (2)
  b. Middle school (3)
  c. University (4)
6. What degree does the father have?
  a. No degree (0)
  b. Junior high school (intermediate school certificate) (1)
  c. Secondary school (2)
  d. Middle school (3)
  e. University (4)

**Figure A.1.**
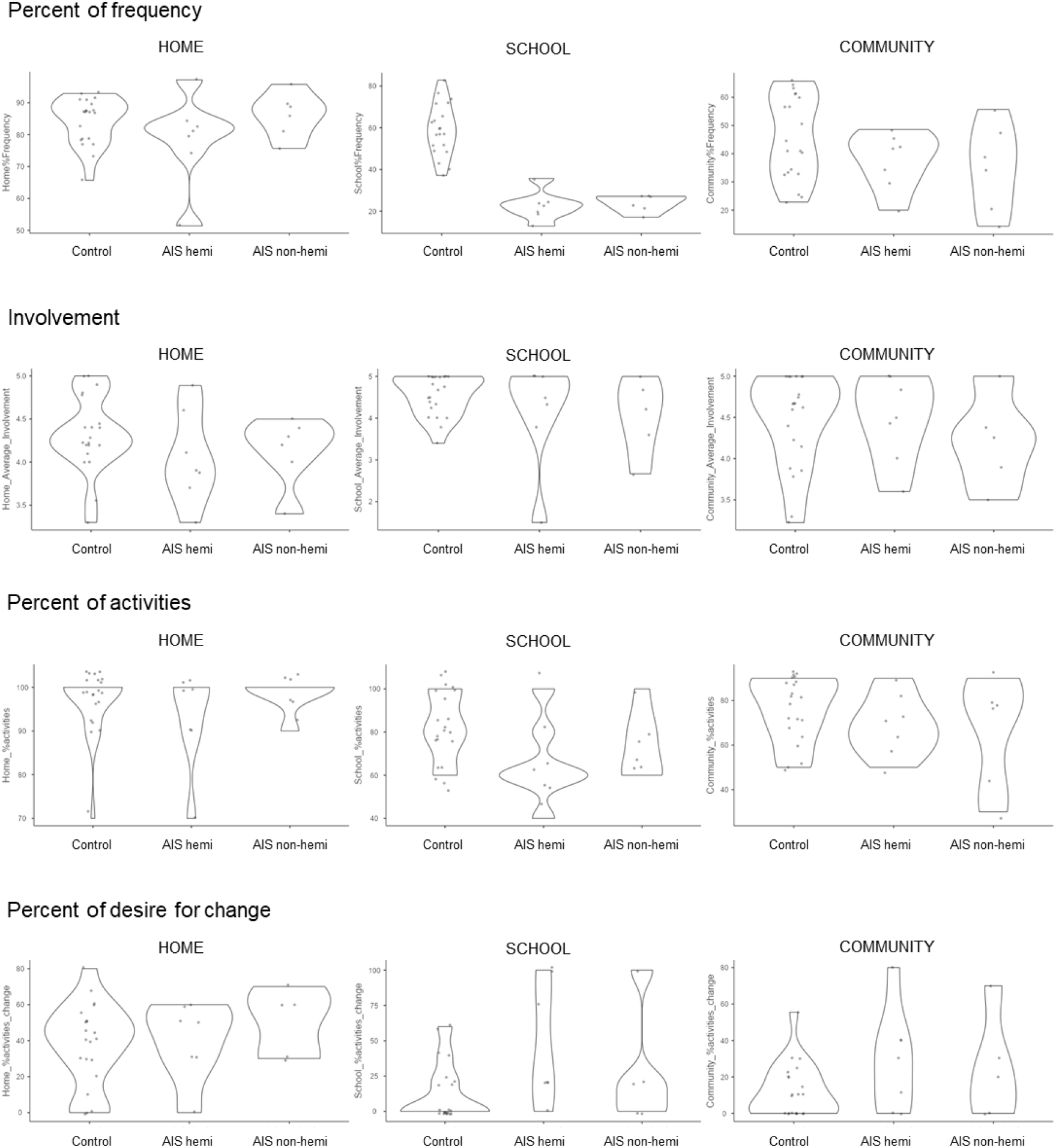
Scatter and violin plots of the participation data (frequency, involvement, activities, and desire for change) derived from the PEM-CY questionnaire in each domain (home, school, and community) in each cohort (control, AIS with hemiparesis and AIS without hemiparesis).

**Table A.1.** Relation between hand strength asymmetry and participation based on the zero-order bivariate correlations and partial correlation (controlling for lesion size, ABILHAND-Kids and SES) with Spearman’s rho correlation coefficients, as well as the change in correlation coefficients.

|  |  | Hemiparesis severity (Hand strength asymmetry) |  |  |  |  |
| --- | --- | --- | --- | --- | --- | --- |
|  |  | Zero-order bivariate correlation |  | Partial correlation (lesion size, ABILHAND-Kids and SES) |  | Change in correlation coefficient from zero-order to partial correlations |
|  |  | rho | interpretation | rho | interpretation |  |
| **Participation at home** | % Frequency | -0.40 | Weak | -0.51 | Moderate | ▲ |
|  | Involvement | -0.23 | None | -0.64<sup>‡</sup> | Moderate | ▲ |
|  | % of activities | -0.08 | None | 0.11 | None | ▬ |
|  | % of desire for change | 0.16 | None | 0.16 | None | ▬ |
| **Participation at school** | % Frequency | -0.47 | Weak | -0.13 | None | ▼ |
|  | Involvement | -0.02 | None | 0.02 | None | ▬ |
|  | % of activities | -0.54<sup>‡</sup> | Moderate | -0.21 | None | ▼ |
|  | % of desire for change | 0.55<sup>‡</sup> | Moderate | 0.43 | Weak | ▼ |
| **Participation in the community** | % Frequency | -0.05 | None | 0.17 | None | ▬ |
|  | Involvement | -0.09 | None | -0.54 | Moderate | ▲ |
|  | % of activities | -0.37 | Weak | 0.03 | None | ▼ |
|  | % of desire for change | 0.39 | Weak | 0.53 | Moderate | ▲ |

The icons represent how the interpretation of the correlation coefficients changed (▬ no change, ▲ increase in the correlation coefficient, and ▼ decrease in the correlation coefficient) from the simple bivariate correlation to the partial correlation, controlling for the indicated variables. If no change, the covariates held constant had little influence on the studied relationship, whilst if there was either an increase or decrease, the variables had an influence between hand strength asymmetry and participation. <sup>‡</sup> Trend toward significant correlation ( $p < 0.07$ ).

